# Adapting Echocardiographic Foundation Models for Detecting Pulmonary Embolism-Related Right-Heart Strain

**DOI:** 10.64898/2026.11.20.26368600

**Authors:** Luming Yang, Yasemin Ozkut, Zhenyu Bu, Siqing Li, Chang Yu, Elaine Situ-LaCasse, Josie Acuña, Adrienne Yarnish, Martin Demant, Srikar R. Adhikari, Alper Yilmaz

## Abstract

Pulmonary embolism (PE) is a time-sensitive medical emergency in which rapid recognition of right ventricular (RV) dysfunction by point-of-care ultrasound (POCUS) can support early risk stratification. Here, we present HOPE (ecHO–Pulmonary Embolism), a real-world emergency cardiac POCUS video dataset comprising 4,069 patients with suspected PE, and EchoPE^1^, a foundation-model-based video classification framework for detecting PE-associated right-heart phenotypes. HOPE includes five commonly acquired emergency echocardiographic views, with all positive PE cases confirmed by computed tomography pulmonary angiography. EchoPE builds on EchoPrime-pretrained echocardiographic representations and incorporates view recognition, task-specific adaptation, and attention-based interpretability. Experiments suggested that directly transferring EchoPrime representations to this disease-specific task provided limited performance, whereas EchoPE (AUROC 95.2 ± .3) substantially outperformed both EchoPrime with a classification head (AUROC 87.6 ± 1) and conventional video deep learning baselines. At a fixed sensitivity of 90%, EchoPE achieved a precision of 87.8, compared with 70.7 for EchoPrime. Among adaptation strategies, Low-Rank Adaptation combined with knowledge distillation provided the best balance between PE discrimination and preservation of pretrained echocardiographic semantics. Interpretability analyses showed that the adapted model focused on clinically relevant RV morphological changes, interventricular septal configuration, and informative temporal segments within ultrasound videos. Overall, HOPE and EchoPE demonstrate that echocardiographic foundation models require dataset-specific adaptation to identify PE-related right-heart strain and support the potential use of artificial intelligence-assisted bedside echocardiography for risk warning in acute care settings.

## 1 Main

Pulmonary embolism (PE) is a common, time-sensitive, and potentially life-threatening diagnostic challenge in the emergency department. In the United States, venous thromboembolism (VTE), which includes deep vein thrombosis and PE, affects up to 900,000 people annually and is associated with more than 60,000 deaths estimated each year [1]. National inpatient data further suggest that PE-related hospital admissions have increased substantially over time, rising from 23 to 65 admissions per 100,000 population between 1993 and 2012 [2]. Despite this burden, early diagnosis remains difficult because PE symptoms are often nonspecific and overlap with other acute cardiopulmonary conditions. A systematic review found that patients may have symptoms for nearly one week before PE is diagnosed, with more than seven days of delay occurring in approximately one-quarter of patients [3]. This delay is clinically consequential because the major mechanism of early morbidity and death in severe PE is acute right ventricular (RV) pressure overload, RV dysfunction, and subsequent hemodynamic collapse. Patients with massive PE, defined by systolic arterial pressure below 90 mmHg, had a 90-day mortality rate of 52.4%, compared with 14.7% among patients without massive PE [4]. Accordingly, risk stratification in PE depends not only on hemodynamic status but also on rapid assessment of RV pressure overload and RV function [5, 6].

Current guidelines identify computed tomography pulmonary angiography (CTPA) as a key imaging modality for confirming PE [7]. However, CTPA may be delayed or infeasible in patients who are hemodynamically unstable, difficult to transport, or being evaluated in resource-limited acute-care environments. In these settings, point-of-care ultrasound (POCUS) can rapidly evaluate signs of PE, including RV dilatation, abnormal interventricular septal motion, McConnell’s sign [8], RV dysfunction, *etc.*, thereby supporting early recognition and treatment prioritization. Emergency physician-performed bedside echocardiography has shown high specificity for PE [9], and systematic reviews indicate that several echocardiographic and multiorgan POCUS findings are highly specific rule-in signs for PE [10, 11]. Focused cardiac ultrasound has therefore become an important bedside tool for expediting diagnostic evaluation and triage in emergent settings [12]. Nevertheless, the acquisition and interpretation of echocardiographic images remain operator dependent, and accurate recognition of RV-related PE phenotypes requires training that is not uniformly available across non-academic, community, and resource-limited settings. These limitations motivate the development of automated, AIenabled echocardiographic tools that can assist clinicians with limited ultrasound expertise in detecting PE-related RV pressure-overload patterns at the point of care.

In recent years, deep learning has substantially advanced automated echocardiographic analysis [13–15]. Early studies demonstrated that deep learning models can perform standard echocardiographic view classification and achieve expert-level or near expert-level performance across multiple transthoracic echocardiography views [16,17]. More broadly, medical imaging foundation models, segmentation networks, and visual pretraining methods have developed rapidly [18–20]. EchoNet-Dynamic further showed that video-based models can automatically segment the left ventricle, estimate ejection fraction, and identify systolic dysfunction from echocardiographic videos [21]. In addition, AI systems have been evaluated for assisting in cardiac function assessment and video-quality evaluation in emergency department POCUS, suggesting the potential for deep learning models to be translated into bedside, low-resource, and operator-variable clinical environments [22,23].

Despite these advances, most existing AI studies in echocardiography have focused on view recognition, left ventricular ejection fraction, structural heart disease, or comprehensive transthoracic echocardiography report generation [15, 21, 24, 25]. Public datasets and model studies specifically targeting PE or RV pressure-overload phenotypes in emergency POCUS remain scarce, and disease-specific modeling for this setting is still underdeveloped [26]. In particular, it remains unclear how general-purpose pretrained echocardiographic models can be adapted to a specific, high-risk, RV-related acute-care task [27]. Conventional task-specific models typically require extensive manual annotation and often show limited generalizability in small-sample settings, low-quality videos, non-standard views, and cross-institutional deployment [28–30]. Moreover, the reported diagnostic performance of deep learning models in medical imaging may be overestimated when study design, validation, and reporting standards are insufficiently rigorous [28, 29]. Recent echocardiography foundation models offer a promising alternative. EchoCLIP learns general echocardiographic representations from paired cardiac ultrasound images and expert text reports [24]. EchoPrime further leverages large-scale video–report pairs, view-aware modeling, multi-video aggregation, and anatomical attention to support a comprehensive interpretation of full echocardiographic examinations, with advantages in low-resource and low-quality imaging scenarios [25]. Nevertheless, the ability of such models to analyze RV-related phenotypes remains limited, particularly in acute conditions where RV function is closely linked to morbidity and mortality [25, 31].

Against this background, we introduce the HOPE dataset and the EchoPE framework to explore a clinically oriented pathway for translation: leveraging the view-recognition and video-representation capabilities of existing echocardiography foundation models to learn PE-related RV information from real-world emergency POCUS data, and evaluating clinical utility through fine-tuning strategies and interpretability analyses.

Specifically, we constructed HOPE, a real-world emergency POCUS echocardiogram video dataset of suspected PE cases, comprising 4,069 patients and five commonly used bedside cardiac ultrasound views. Unlike conventional imaging datasets centered on CTPA, HOPE reflects the clinical context in which emergency physicians make time-sensitive bedside decisions based on cardiac ultrasound findings. It therefore provides an opportunity to evaluate the potential of AI models for emergency screening, risk signaling, and recognition of RV pressure overload [5, 27].

Using this dataset, we developed EchoPE, a video-classification framework for PE-related echocardiographic assessment. EchoPE leverages representations extracted by pretrained echocardiographic models and applies them to both view recognition and ultrasound video classification. Rather than training task-specific models from scratch, EchoPE investigates how large pretrained cardiac ultrasound models can be transferred to an underrepresented but clinically important RV-related disease phenotype. We systematically compared multiple adaptation strategies, including frozen-feature training, low-rank adaptation (LoRA), partial fine-tuning, and full fine-tuning, and benchmarked their performance against representative 3D convolutional neural network and Transformer-based baselines.

Finally, we performed attention-based interpretability analyses to link model predictions with PE-related RV pressure-overload phenotypes and to examine the contribution of different video segments and anatomical regions to classification. Through HOPE and EchoPE, this study presents an end-to-end workflow spanning real-world emergency data curation, pretrained model adaptation, RV-related disease-phenotype learning, and interpretability assessment. Together, these components provide a generalizable framework for improving the clinical usability of pretrained echocardiographic models in acute-care POCUS.

## 2 Results

### 2.1 Clinical cohort and characteristics of the HOPE dataset

We constructed the HOPE dataset (*ecHO-Pulmonary Embolism*), a real-world emergency department point-of-care ultrasound (POCUS) video dataset designed to support the automated recognition of pulmonary embolism (PE)-related right-heart strain phenotypes. The dataset included patients who underwent POCUS in the emergency department due to clinical suspicion of PE or the presence of PE-related risk features. HOPE represents the first large-scale, real-world emergency department POCUS video dataset specifically curated for the automated recognition of PE-related right-heart strain phenotypes.

HOPE contains 4,069 POCUS videos that were annotated by emergency physicians according to ultrasound findings in two categories: *normal* and *PE-related right-heart phenotype*. The normal group comprised videos without sonographic evidence of PE-related right-heart overload or dysfunction. The PE-related right-heart phenotype group comprised videos demonstrating imaging features consistent with right-sided pressure or volume overload in the context of PE, including right ventricular enlargement, an abnormal right-to-left ventricular size relationship, interventricular septal flattening or paradoxical septal motion, impaired right ventricular systolic function, findings consistent with McConnell’s sign, and other right-heart abnormalities considered compatible with PE-associated cardiac involvement [32].

The overall class distribution was approximately balanced, with 1,934 normal videos and 2,135 videos labeled as PE-related right-heart phenotype, corresponding to a normal-to-PE ratio of approximately 1:1.104. All patients labeled as having a PE-related right-heart phenotype had PE confirmed by CTPA, and conversely, all patients labeled as negative had no evidence of PE or other abnormalities on CTPA.

The dataset covered five commonly acquired emergency POCUS cardiac or hemodynamic views: apical four-chamber view (A4), parasternal short-axis view (PSS), parasternal long-axis view (PSL), inferior vena cava view (IVC), and subxiphoid view (SX). For A4-view videos, expert annotators additionally provided labels for the presence or absence of McConnell’s sign. The distribution of samples across views and the distribution of video frame counts are shown in Fig. 1a. Additional dataset characteristics are provided in the Supplementary Information.

**Figure 1.**
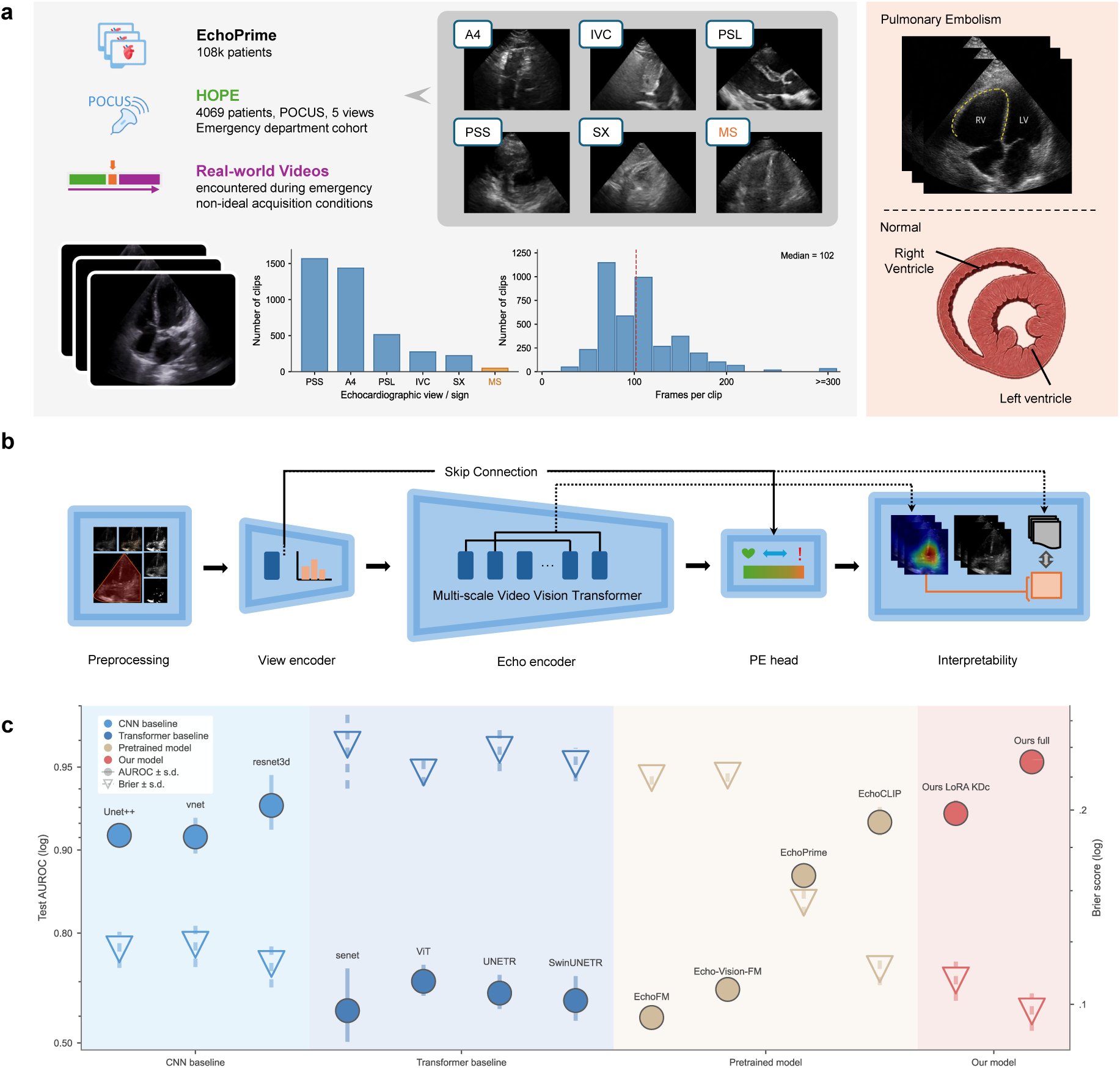
Overview of the HOPE Dataset and EchoPE Framework. a, Dataset composition; b, EchoPE workflow; and c, cross-validation performance across models. **(a)** Distribution of echocardiographic views and clip lengths in the study cohort. Left, labeled view distribution across 4,069 clips. Right, per-clip frame-count histogram using 20-frame bins from 0 to 300 frames, with a final bin for clips containing more than 300 frames; the red dashed line denotes the median clip length of 102 frames. Compared right-versus left-ventricular representation and Normal versus pulmonary embolism (PE) studies (orange box). **(b)** Overview of the EchoPE pipeline. **(c)** Five-fold cross-validation performance across model families. Filled circles show test area under the receiver operating characteristic curve (AUROC; left axis), and open downward triangles show Brier score (right axis; lower is better). Error bars indicate standard deviation across folds, and shaded regions denote model families.

HOPE captures substantial real-world variability commonly encountered during emergency bedside ultrasound. Videos include non-ideal acquisition conditions such as off-axis views, incomplete visualization of the cardiac sector, suboptimal gain or depth settings, motion artifacts, respiratory motion, probe compression, limited acoustic windows, and heterogeneous image quality. This heterogeneity reflects the operational complexity of emergency POCUS and provides a clinically relevant benchmark for evaluating the robustness and transferability of video-based deep learning models under resource-constrained, time-sensitive, and non-standard imaging conditions.

For model initialization, we used the pretrained weights of EchoPrime which was pretrained on 12 million cardiac ultrasound videos and reports from 108k patients, spanning multiple large-scale datasets. These included datasets from CSMC, SHC, MIMIC, CGMH, and other predominantly private institutional sources [25].

### 2.2 Overview of the EchoPE framework

Using the HOPE dataset, we developed EchoPE, a video classification framework for recognizing PE-related right-heart ultrasound phenotypes. The primary goal of EchoPE was not to train a task-specific model from scratch, but to adapt a large-scale pretrained echocardiogram foundation model to a clinically important POCUS task. EchoPE consists of five major components: a preprocessing module, a view encoder, an echo encoder, a PE classification head, and an interpretability module. The overall workflow is shown in Fig. 1b.

In the preprocessing module, raw MP4 videos were first processed to remove machine interface information, segment the ultrasound sector, crop the spatial region of interest, and standardize the video size. The resulting purified video sequence had a fixed spatial resolution of 256 × 256. During training, data augmentation was subsequently applied. This preprocessing step was designed to reduce the influence of non-medical regions, including device interface elements, textual overlays, and electrocardiographic traces, while also harmonizing videos acquired from different ultrasound systems and operators. This step is particularly important for reducing the risk of shortcut learning. The effect of preprocessing on model performance was evaluated in supplementary experiments.

The view encoder was used to identify the echocardiogram view of the input video. Unlike the video-level echo encoder, the view encoder performed image classification using only the first frame of each video, thereby reducing computational cost while providing a rapid view-level prior. The predicted view embedding was passed to the PE classification head through a skip connection and combined with the video-level representation generated by the echo encoder. This design allowed the model to adaptively learn PE-related ultrasound features across different views and improved the interpretability of the model outputs.

The echo encoder formed the core video representation module of EchoPE and was initialized using EchoPrime pretrained weights. In EchoPE, the echo encoder received preprocessed video clips and used spatiotemporal attention mechanisms to learn information related to cardiac motion, right ventricular morphology, dynamic septal abnormalities, and IVC motion, all of which may be relevant to PE-associated right-heart strain.

The PE classification head integrated the video-level features from the echo encoder and the view-level features from the view encoder to output the probability of *normal* versus *PE-related right-heart phenotype*. The interpretability module was used to analyze the basis of model predictions. This module generated explanatory outputs based on attention maps, temporal attention, and frame-level attribution, highlighting the temporal segments and anatomical regions most influential for decisions. EchoPE therefore outputs an auxiliary report containing the predicted probability, predicted view, key frames, regions of attention, and explanations.

### 2.3 Task-specific fine-tuning substantially improved PE phenotype recognition

We first evaluated the direct transferability of a pretrained cardiac ultrasound model to the HOPE dataset without task-specific fine-tuning. The original EchoPrime model can generate comprehensive representations and reports from full echocardiographic examinations; however, its performance was limited when directly applied to the binary classification of PE-related right-heart phenotypes in HOPE. Using the original EchoPrime representations with prototype learning or zero-shot classification, the model achieved an AUROC of 0.571 for right ventricular systolic function, 0.535 for right ventricular dilation, and 0.612 for pulmonary artery pressure in the internal evaluation set (Fig. 2a).

**Figure 2.**
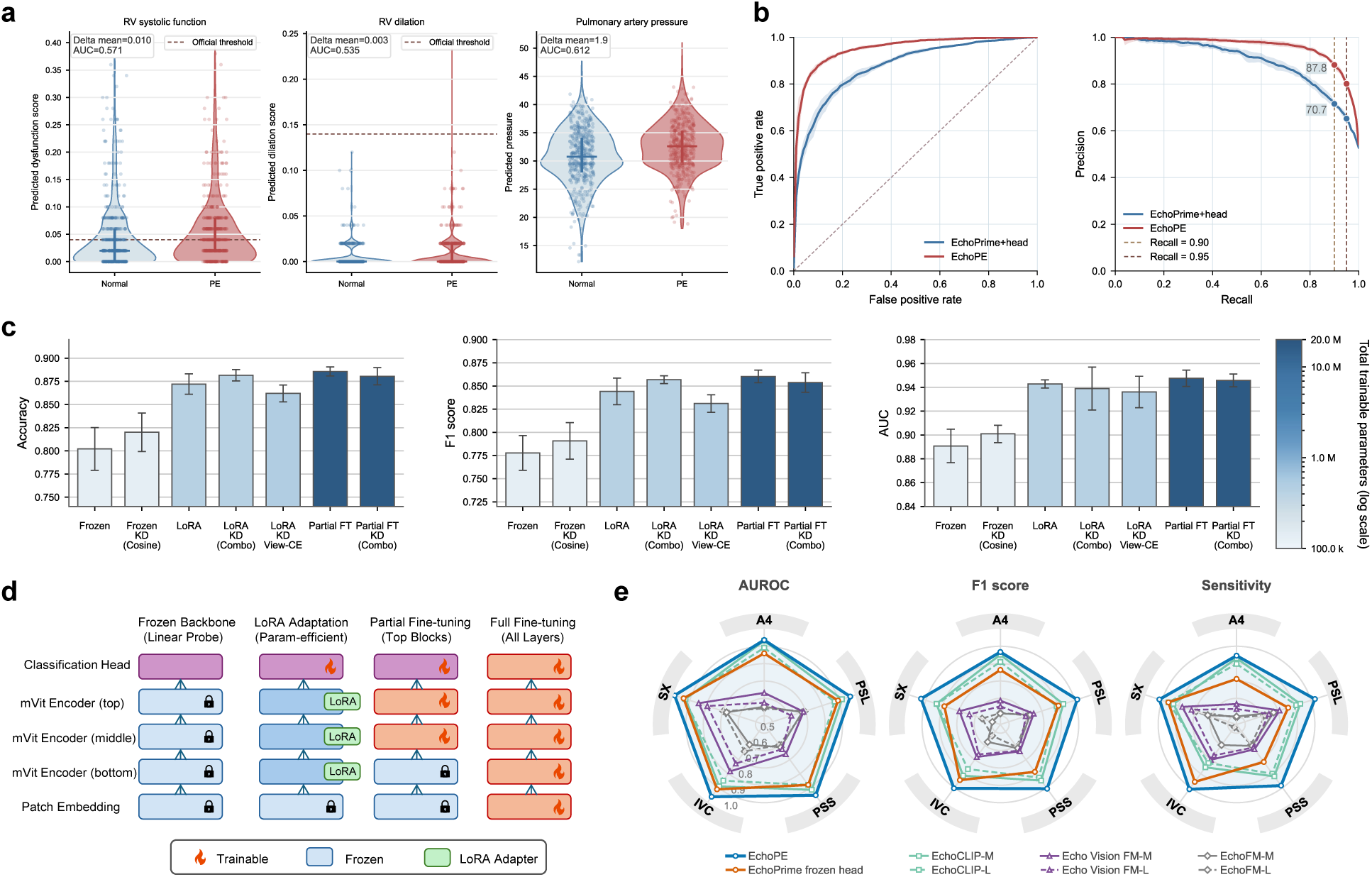
Comparative Experiment for EchoPE. **(a)** EchoPrime-derived phenotype predictions in Normal and PE videos for right-ventricular systolic function, right-ventricular dilation and pulmonary artery pressure. Violin plots with jittered observations show per-video predictions. **(b)** Pooled five-fold test-set ROC and precision–recall curves comparing EchoPrime+head with EchoPE. **(c)** Cross-validation comparison of seven fine-tuning configurations ordered by increasing trainable parameter count: frozen backbone, frozen + knowledge distillation (KD), LoRA, LoRA + KD, LoRA + KD + view-level cross-entropy, partial fine-tuning, and partial fine-tuning + KD. Bars show mean ± s.d.; color intensity encodes trainable parameter count on a logarithmic scale. **(d)** Schematic comparison of the fine-tuning strategies evaluated in c. **(e)** View-stratified radar plots showing mean AUROC, F1 score and sensitivity across five standard views: A4, PSL, PSS, IVC and SX. Solid and dashed lines denote MLP and linear heads, respectively; the shaded polygon highlights full EchoPE fine-tuning.

We then evaluated EchoPE and a baseline consisting of EchoPrime features with an additional classification head. EchoPE achieved an AUROC of 0.9521 ± 0.0031, whereas EchoPrime with a classification head achieved an AUROC of 0.8763 ± 0.0097. These results indicate that the representations learned from large-scale cardiac ultrasound pretraining provide useful prior context for recognizing PE-related right-heart phenotypes, but that task-specific adaptation and architectural components designed for emergency POCUS substantially improve performance.

To assess clinical usability at high-sensitivity operating points, we fixed recall (sensitivity) at 0.90 and 0.95 and compared the resulting precision. At a fixed recall of 0.90, EchoPE achieved a precision of 0.8778, whereas EchoPrime with a classification head achieved a precision of 0.7067. At a fixed recall of 0.95, the corresponding precisions were 0.7960 and 0.6487, respectively (the Supplementary Information). These findings suggest that, at clinically relevant high-sensitivity thresholds, EchoPE substantially improved precision and thereby reduced the proportion of false positives among flagged cases.

### 2.4 Comparison with baseline architectures and pretrained foundation models

We compared our method with conventional deep learning architectures and pretrained cardiac ultrasound foundation models using the same preprocessing pipeline, evaluation metrics, and 5-fold cross-validation splits. As shown in Table 1, all reported results are presented as mean ± standard deviation across the five folds.

**Table 1.**
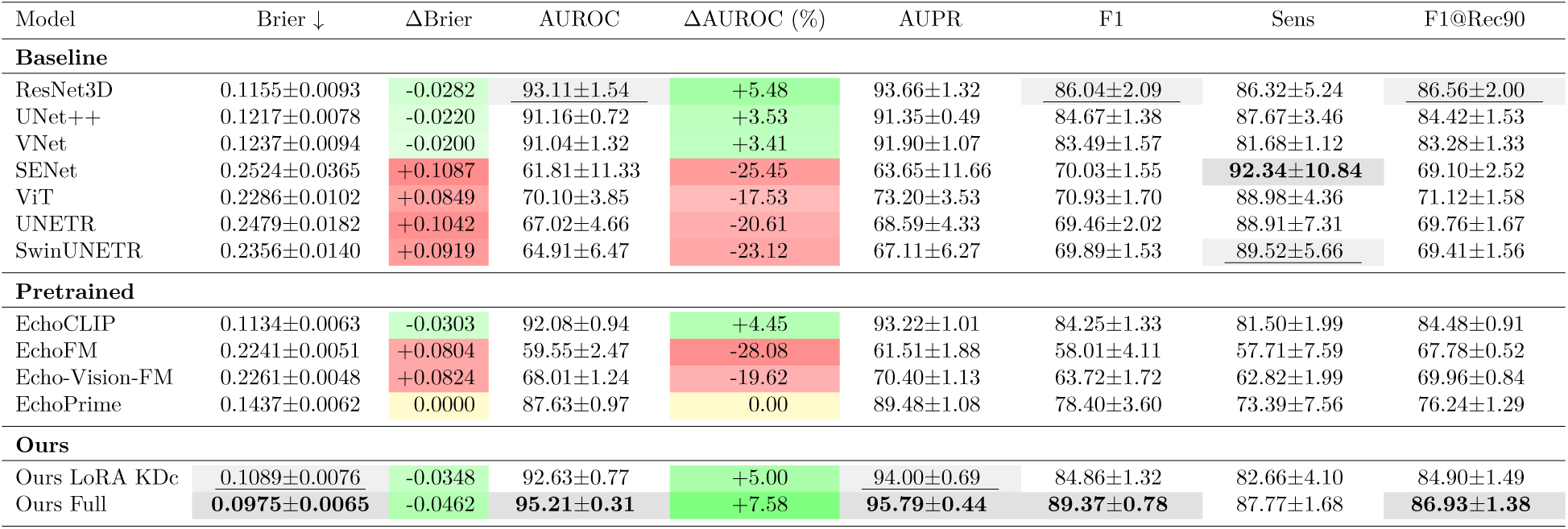
Performance comparison among different models. All metrics except Brier are reported in percentage (%). Δ indicates change relative to EchoPrime. Results include mean ± std. Green indicates improvement over EchoPrime (Brier ↓, AUROC ↑), Red indicates performance degradation, and **bold** / <u>underline</u> denote the best and second-best results, respectively.

| Model | Brier ↓ | ΔBrier | AUROC | ΔAUROC (%) | AUPR | F1 | Sens | F1@Rec90 |
|---|---|---|---|---|---|---|---|---|
| **Baseline** |  |  |  |  |  |  |  |  |
| ResNet3D | 0.1155±0.0093 | -0.0282 | 93.11±1.54 | +5.48 | 93.66±1.32 | 86.04±2.09 | 86.32±5.24 | 86.56±2.00 |
| UNet++ | 0.1217±0.0078 | -0.0220 | 91.16±0.72 | +3.53 | 91.35±0.49 | 84.67±1.38 | 87.67±3.46 | 84.42±1.53 |
| VNet | 0.1237±0.0094 | -0.0200 | 91.04±1.32 | +3.41 | 91.90±1.07 | 83.49±1.57 | 81.68±1.12 | 83.28±1.33 |
| SENet | 0.2524±0.0365 | +0.1087 | 61.81±11.33 | -25.45 | 63.65±11.66 | 70.03±1.55 | **92.34±10.84** | 69.10±2.52 |
| ViT | 0.2286±0.0102 | +0.0849 | 70.10±3.85 | -17.53 | 73.20±3.53 | 70.93±1.70 | 88.98±4.36 | 71.12±1.58 |
| UNETR | 0.2479±0.0182 | +0.1042 | 67.02±4.66 | -20.61 | 68.59±4.33 | 69.46±2.02 | 88.91±7.31 | 69.76±1.67 |
| SwinUNETR | 0.2356±0.0140 | +0.0919 | 64.91±6.47 | -23.12 | 67.11±6.27 | 69.89±1.53 | 89.52±5.66 | 69.41±1.56 |
| **Pretrained** |  |  |  |  |  |  |  |  |
| EchoCLIP | 0.1134±0.0063 | -0.0303 | 92.08±0.94 | +4.45 | 93.22±1.01 | 84.25±1.33 | 81.50±1.99 | 84.48±0.91 |
| EchoFM | 0.2241±0.0051 | +0.0804 | 59.55±2.47 | -28.08 | 61.51±1.88 | 58.01±4.11 | 57.71±7.59 | 67.78±0.52 |
| Echo-Vision-FM | 0.2261±0.0048 | +0.0824 | 68.01±1.24 | -19.62 | 70.40±1.13 | 63.72±1.72 | 62.82±1.99 | 69.96±0.84 |
| EchoPrime | 0.1437±0.0062 | 0.0000 | 87.63±0.97 | 0.00 | 89.48±1.08 | 78.40±3.60 | 73.39±7.56 | 76.24±1.29 |
| **Ours** |  |  |  |  |  |  |  |  |
| Ours LoRA KDc | 0.1089±0.0076 | -0.0348 | 92.63±0.77 | +5.00 | 94.00±0.69 | 84.86±1.32 | 82.66±4.10 | 84.90±1.49 |
| Ours Full | **0.0975±0.0065** | -0.0462 | **95.21±0.31** | +7.58 | **95.79±0.44** | **89.37±0.78** | 87.77±1.68 | **86.93±1.38** |

Among conventional models, ResNet3D [33] achieved the strongest overall performance, with an AUROC of 93.11 ± 1.54, outperforming other non-pretrained architectures. UNet++ [34] and VNet [35] also showed reasonable discrimination, whereas SENet [36], ViT [37], UNETR [38], and SwinUNETR [39] performed substantially worse in terms of AUROC and calibration. Notably, several transformer-based models achieved relatively high sensitivity despite low AUROC and F1 scores. This pattern arguably does not reflect robust discriminative ability, as high sensitivity was accompanied by poorer overall ranking performance and reduced precision-sensitive metrics.

Pretrained cardiac ultrasound foundation models showed competitive performance while requiring substantially fewer trainable parameters, as only the MLP classification head was fine-tuned. See details in the Supplementary Information. EchoCLIP [24] was the strongest among these models, including EchoFM [40] and Echo-Vision-FM [31], achieving an AUROC of 92.08 ± 0.94.

Our proposed models consistently improved upon the EchoPrime reference model. Compared with the strongest baseline, EchoPE further improved AUROC by 2.10 percentage points.

These results suggest that PE-related right-heart abnormalities can be captured from emergency cardiac ultrasound videos, and that pretrained cardiac ultrasound representations can transfer to PE-related POCUS classification under lightweight fine-tuning. However, effective performance requires both cardiac ultrasound pretraining and PE-specific video-level adaptation.

### 2.5 Fine-tuning strategies for pretrained cardiac ultrasound models

We next systematically compared multiple adaptation strategies in our ablation setting for pretrained models, including frozen feature training, low-rank adaptation (LoRA), partial fine-tuning, and proposed variants with knowledge distillation. Schematic illustrations of these strategies are shown in Fig. 2d, and experimental results are shown in Fig. 2c. This ablation was conducted under a separate subset, fixed train/validation/test configuration designed for controlled comparison of adaptation strategies.

Among all adaptation strategies, partial fine-tuning achieved the best overall performance in this ablation setting. Compared with frozen feature training, partial fine-tuning improved the AUROC by 10.4%, reaching 0.8857 ± 0.0049. LoRA provided a favorable trade-off between performance and parameter efficiency, achieving an AUROC of 0.8721 ± 0.0110. Adding knowledge distillation further improved LoRA performance: our proposed LoRA KD-combo strategy achieved an internal test-set AUROC of 0.8816 ± 0.0062, which was comparable to PFT KD-combo (0.8805 ± 0.0093). Compared with standard LoRA, the KD-combo strategy produced more stable performance, suggesting that knowledge distillation may help preserve general pretrained representations while enhancing task-specific adaptation to PE-related phenotypes.

Because PE-related right-heart strain features are differentially visible across POCUS views, we further analyzed classification performance by view. Results are shown in Fig. 2e. Across models, the comparison of multilayer perceptron and linear-probe classification heads showed that the multilayer perceptron head was more effective for this task.

EchoPE achieved high and relatively stable performance across all five views, although performance varied by view. The SX view showed the highest performance, with an AUROC of 0.9912 ± 0.0106, suggesting that subxiphoid imaging may provide highly discriminative information related to right-heart strain in this dataset. PSL, IVC, and PSS views also achieved strong performance, with AUROCs of 0.9699 ± 0.0094, 0.9643±0.0192, and 0.9534±0.0048, respectively. PSL and PSS views can capture right ventricular morphology and interventricular septal motion, whereas the IVC view may provide indirect information related to volume status and right-sided pressure. The A4 view achieved an AUROC of 0.9360 ± 0.0125. Although slightly lower than the other views, this performance remained high, indicating that right ventricular enlargement and systolic dysfunction in the apical four-chamber view are also important features for identifying PE-related right-heart phenotypes.

Overall, these findings support the ability of EchoPE to recognize PE-related right-heart phenotypes across multiple commonly acquired emergency POCUS views.

### 2.6 View analysis

A central objective of the view analysis was to evaluate the robustness of view classification because accurate view assignment is critical for downstream pulmonary embolism (PE) prediction. The dataset contained five echocardiographic views: apical four-chamber (A4), inferior vena cava (IVC), parasternal short-axis (PSS), parasternal long-axis (PSL), and subxiphoid (SX). In addition, A4 videos with McConnell’s sign were annotated separately as MS, representing a PE-associated imaging phenotype. We evaluated view classification under three label granularities: the original six-class setting, consisting of five views plus MS; a four-class setting, in which A4 and MS were merged into A4, and IVC and SX were merged into a subxiphoid category; and a three-class setting, in which PSL and PSS were further merged into a parasternal category.

We first examined how the proportion of HOPE training data affected model performance across different training strategies, including the direct use of the pretrained model, the pretrained model with linear probing, the pretrained model with full fine-tuning, the pretrained model with partial fine-tuning, and training from scratch (Fig. 3a). Because the EchoPrime pretrained model was not trained on point-of-care ultrasound views and its native view taxonomy did not include SX or MS, the direct pretrained evaluation was restricted to the aggregated labels shared with the available EchoPrime view outputs.

**Figure 3.**
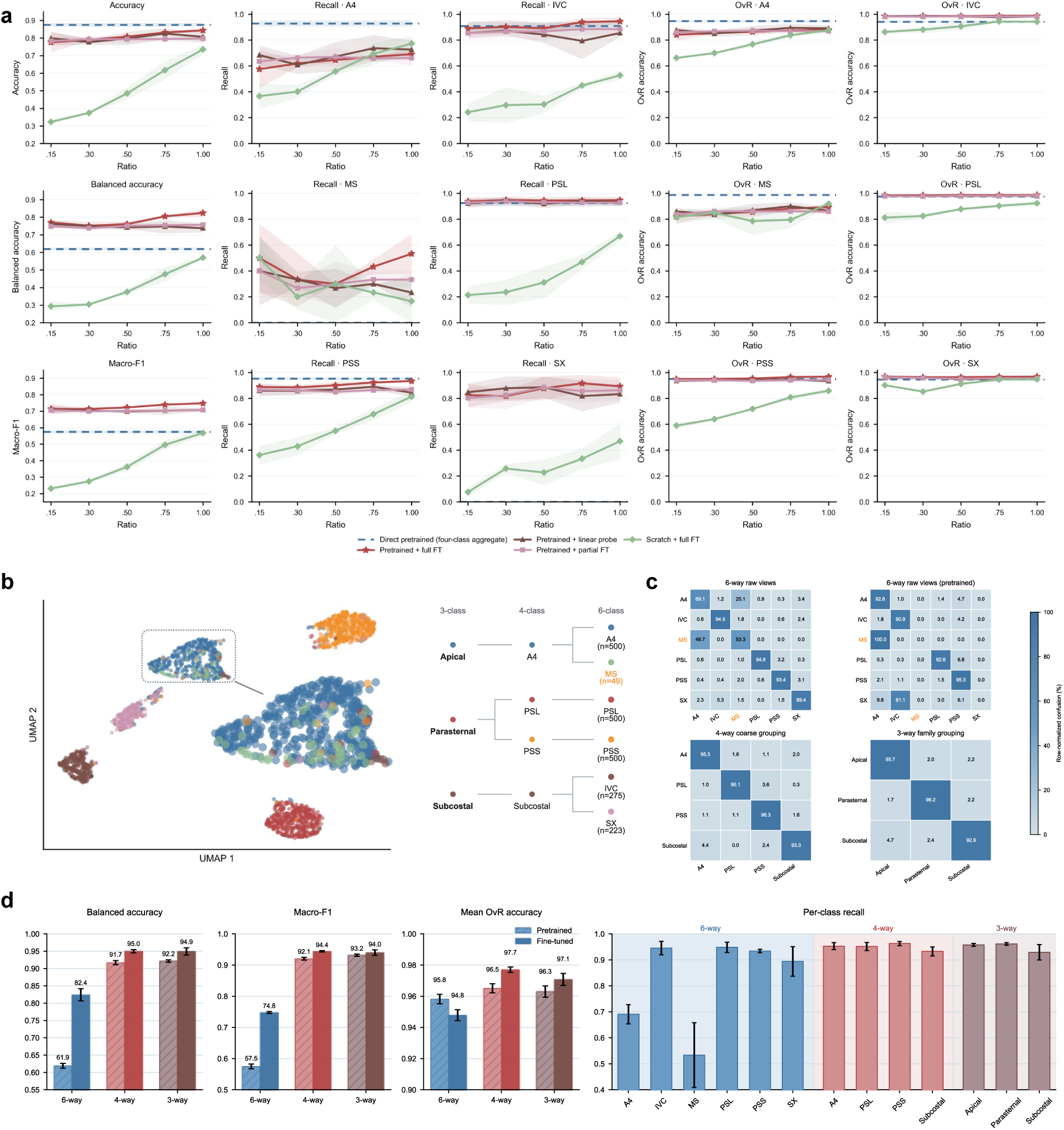
Experimental Result for Echocardiographic View Classification. **(a)** Effect of training-set size on view-classification transfer. Overall metrics, per-class recall and one-versus-rest accuracy are shown across training subset ratios. Overall accuracy, balanced accuracy and macro-F1 are reported as mean ± s.d. across random seeds. Direct pretrained inference is shown as a horizontal dashed baseline for aggregate four-way accuracy. **(b)** Hidden-space organization of the final EchoPrime view classifier. Penultimate ConvNeXt features from the six-way pretrained full fine-tuned model were projected with UMAP. Points are colored by six raw view labels. The projection was fit on a class-capped subset of 2,047 videos, with up to 500 examples per raw view. **(c)**, Row-normalized confusion matrices for deployment models at the full training-set ratio. Percentages were computed from the mean confusion matrix across seeds. **(d)** Label-space summary comparing pretrained and full fine-tuned EchoPrime view classifiers. Overall metrics are shown together with per-class recall for the fine-tuned deployment model.

Overall, view classification was highly data-dependent. Training from scratch with 4,069 HOPE examples was insufficient to obtain consistently strong performance. In contrast, strategies initialized from pretrained weights achieved substantially better results. Linear probing, partial fine-tuning, and full fine-tuning all performed well, indicating that pretrained representations provided a strong basis for view recognition. Among these approaches, partial fine-tuning achieved the best overall performance, outperforming full fine-tuning. Partial fine-tuning was also the most responsive to increases in the proportion of HOPE training data, whereas linear probing and full fine-tuning showed a tendency toward reduced performance at higher data ratios.

The UMAP visualization of learned embeddings showed clear separation among most view categories (Fig. 3b). A4 and MS strongly overlapped and were difficult to distinguish in embedding space. Given that MS represents a pathological sign observed within the A4 view rather than an anatomically distinct view, this overlap is expected and supports the rationale for merging MS with A4 in the aggregated view taxonomy.

Confusion matrices (Fig. 3c), balanced accuracy, macro F1 score, mean one-vs-rest accuracy, and per-class recall (Fig. 3d) further confirm that A4 and MS were the dominant sources of confusion in the original six-class setting. The four-class setting substantially improved classification performance, while the three-class setting achieved the highest aggregate metrics. However, after merging into three classes, the per-class recall of the subxiphoid category became less stable. Therefore, the four-class setting provided the most favorable compromise: it preserved clinically meaningful view information while maximizing per-class recall and the practical benefits of view classification.

Finally, we performed exploratory experiments, as shown in the Supplementary Information, to test whether explicit cross-view information could improve single-view classification. None of the strategies, including the separated encoder and trained pseudo-view representation network, consistently improved performance. Therefore, cross-view information was not introduced directly into the video encoder. Instead, it was incorporated through skip connections at the final classification and interpretability stages.

### 2.7 Interpretability analysis

To evaluate how different adaptation strategies affected model reasoning and the preservation of pretrained semantics, we compared four models: a ResNet3D trained from scratch, a frozen EchoPrime representation model, a LoRA+KD model, and a fully fine-tuned model. Fig. 4a shows UMAP embeddings, spatial attribution maps, attention maps, gradient × input maps, temporal occlusion sensitivity, and scaled attention across video frames. Fig. 4b summarizes the LoRA+KD training framework. Fig. 4c quantitatively compares the models in terms of ROI attribution, semantic preservation, and foreground occlusion experiments.

**Figure 4.**
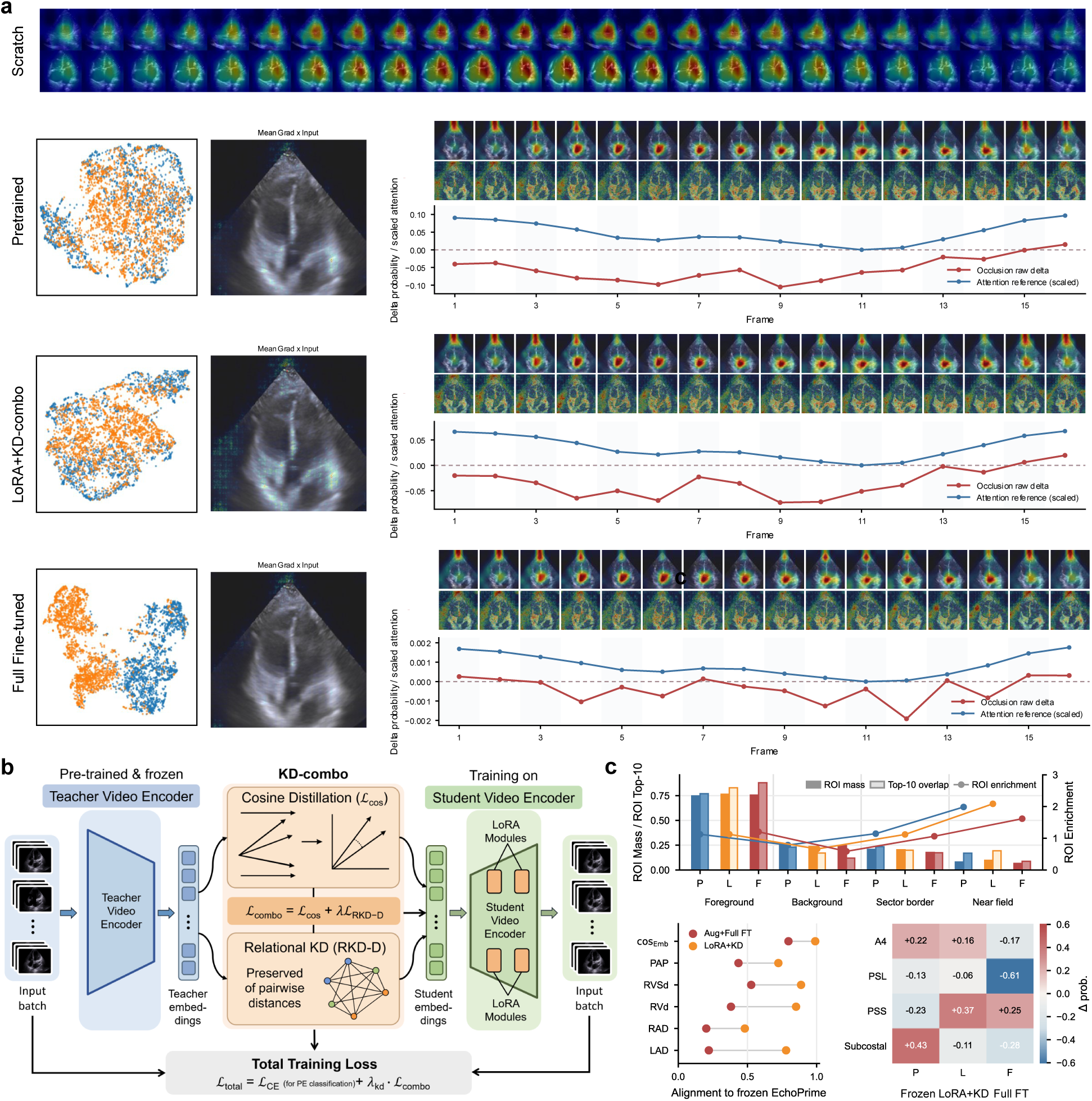
Interpretability Across Fine-Tuning Strategies. **(a)** Case-based interpretability for a representative A4-view test study under three PE-head settings: frozen head, LoRA + KD and full fine-tuning. For each model, the top row shows frame-wise attention-reference overlays, the middle row shows Grad × Input overlays, and the bottom trace shows occlusion delta probability together with a scaled attention-reference signal; the dashed horizontal line denotes zero probability change. Companion panels report clip-averaged Mean Grad × Input overlays. Two-dimensional UMAP projections of 512-dimensional PE-classifier video embeddings are also shown for the frozen head, LoRA + KD and augmented full fine-tuning models. Points are displayed by clinical label, coarse ultrasound view or joint label–view encoding. **(b)** Schematic of the LoRA + KD combination strategy used to adapt EchoPrime representations while preserving semantic alignment. **(c)** Acquisition-context attribution, semantic alignment and foreground-occlusion analysis across PE-head settings. Top, weakly supervised region-of-interest attribution summaries across foreground, background, sector border and probe near-field regions, including ROI mass, top-10 overlap and enrichment. Bottom left, semantic probe alignment between PE-head embeddings, frozen EchoPrime embeddings and phenotype readouts. Bottom right, median foreground-occlusion delta probability stratified by view.

UMAP visualization showed that when the EchoPrime representation was frozen, PE and normal samples still overlapped substantially in embedding space. This indicates that the frozen pretrained representation alone was insufficient for clear PE discrimination in POCUS applications. The LoRA+KD model improved separation between PE and normal samples while largely preserving the global structure of the pretrained representation. In contrast, full fine-tuning further increased class separation but induced a marked reorganization of the embedding space relative to the frozen model, suggesting greater drift from the pretrained semantic space.

Spatial attribution analyzes showed that the ResNet3D trained from scratch primarily focused on the interventricular septal region and areas related to left ventricular compression. The frozen EchoPrime model produced more diffuse attribution patterns, with attention not only to cardiac chambers but also to acquisition-context regions such as the probe near-field and sector border. Both the LoRA+KD and fully fine-tuned models showed more concentrated attribution over PE-relevant cardiac structures, particularly the wall between the right and left ventricles and the relative morphology of the right and left ventricles. Some attention remained distributed over non-cardiac foreground regions.

Temporal analyses showed that scaled attention was biased toward the beginning and end of videos in some samples. In contrast, temporal occlusion experiments showed that removing frames near the middle of the video more often produced larger changes in predicted PE probability. These middle frames may contain more stable information about cardiac chamber morphology and motion, making them more important for final classification. This finding supports the contribution of dynamic video information to PE recognition.

EchoPrime semantic probing showed that LoRA+KD preserved pretrained semantics substantially better than full fine-tuning. The mean and median embedding cosine similarity between LoRA+KD and the frozen EchoPrime model were 0.987 and 0.990, respectively. In comparison, the corresponding values for the fully fine-tuned model decreased to 0.778 and 0.797. Across five internal EchoPrime phenotype readouts, LoRA+KD also maintained stronger agreement with the frozen model. The Spearman correlations for pulmonary artery pressure, right ventricular systolic diameter, right ventricular diameter, right atrial diameter, and left atrial diameter were 0.724, 0.888, 0.852, 0.481, and 0.780, respectively. These results indicate that LoRA+KD can introduce PE-discriminative capacity while preserving the right-heart-related semantic structure encoded in the EchoPrime pretrained representation, whereas full fine-tuning causes more substantial representational reorganization.

ROI attribution analyses further supported this observation. Using four weakly supervised ROI categories— foreground, background, sector border, and probe near-field—the frozen EchoPrime and LoRA+KD models showed highly similar attribution distributions. For the frozen model, the median ROI mass, ROI enrichment, and top-10% overlap were 0.219, 1.139, and 0.292, respectively. For LoRA+KD, the corresponding values were 0.223, 1.132, and 0.303. In contrast, the fully fine-tuned model showed lower corresponding values of 0.192, 1.096, and 0.215. These results suggest that LoRA+KD largely preserved the spatial attribution structure of the frozen EchoPrime model, whereas full fine-tuning altered the attribution distribution.

Finally, occlusion experiments combining the four ROI categories showed that all three EchoPrime-based models were generally robust to the occlusion of any single weakly defined region, with median changes in PE probability close to zero. The median ΔPE probabilities for the frozen EchoPrime, LoRA+KD, and fully fine-tuned models were approximately +7.8 × 10^−5^, −5.2 × 10^−5^, and −1.4 × 10^−6^, respectively. The corresponding median Δlogit values were +0.0058, −0.0567, and −0.131.

Taken together, Fig. 4 shows that LoRA+KD achieved a favorable balance for PE classification adaptation. Compared with the frozen EchoPrime model, it improved separation between PE and normal samples. Compared with full fine-tuning, it substantially reduced semantic drift from the EchoPrime pretrained representation and preserved similar spatial attribution patterns. These findings suggest that LoRA+KD is a suitable adaptation strategy for ultrasound video foundation models, improving task-specific discrimination while retaining clinically relevant semantics from pretraining.

## 3 Discussion

In this study, we constructed the HOPE dataset and proposed the EchoPE framework to identify pulmonary embolism (PE)-related right heart strain phenotypes from real-world emergency point-of-care ultrasound videos. To the best of our knowledge, HOPE is the first clinical dataset designed to identify PE using cardiac ultrasound. Using this dataset, we demonstrate that a pretrained ultrasound foundation model can be effectively adapted to an acute right-heart-related disease-specific clinical task. EchoPE achieved strong discriminative performance in identifying PE-related right heart phenotypes and maintained favorable precision at high-sensitivity operating points, suggesting its potential utility as an emergency screening, risk-alerting, and triage-support tool.

An important finding of this study is that, although a general-purpose pretrained cardiac ultrasound model encodes rich representations of cardiac structure and function, direct transfer to PE-related right heart strain recognition remains limited. Despite the strong performance of the original EchoPrime model in its original training setting, the original EchoPrime representations showed suboptimal zero-shot or prototype-based performance for phenotypes related to right ventricular function, right ventricular enlargement, and pulmonary arterial pressure. This suggests that PE-related POCUS recognition is not simply a matter of reading out generic representations. Insufficient coverage of labels and semantics might be one of the reasons. Furthermore, PE-related right heart strain involves right ventricular morphology, interventricular septal motion, right ventricular systolic function, inferior vena cava dynamics, and contextual information across multiple views. In emergency POCUS, these features are further affected by image quality, non-standard views, and acquisition noise. Therefore, adaptation to the specific clinical task remains necessary. By incorporating a view encoder, a video-level echo encoder, and a task-specific classification head, EchoPE preserves pretrained knowledge while enhancing the recognition of PE-related right heart phenotypes, substantially outperforming the baseline model that directly combines EchoPrime features with a classifier.

Compared with conventional deep learning models trained from scratch, the advantage of EchoPE should be interpreted in a task-specific manner. For the video-level PE phenotype classification task, the 3D ResNet trained from scratch was able to learn meaningful PE-related signals from the HOPE dataset and achieved performance comparable to the EchoCLIP-plus-MLP baseline in some settings. This result suggests that the HOPE dataset contains learnable spatiotemporal information relevant to PE-related right heart strain, and it would be too strong to claim that task-specific data alone are entirely insufficient for learning useful video-level representations. Instead, the main advantage of EchoPE lies in more effectively adapting pretrained cardiac ultrasound knowledge under small-sample, heterogeneous, and clinically noisy POCUS conditions while improving performance and parameter efficiency relative to naïve frozen-feature transfer or fully scratch training. In contrast, the view-classification task based on the first video frame revealed a clearer limitation of training from scratch: even with more than 4000 images, scratch models performed poorly in recognizing ultrasound views. Pretrained models provide prior knowledge regarding cardiac chamber anatomy, myocardial motion, view-specific characteristics, and ultrasound imaging patterns, thereby improving sample efficiency and potential generalizability in downstream tasks. This observation is consistent with the broader development of medical imaging foundation models, in which general representations are learned through large-scale pretraining and subsequently adapted to specific clinical tasks through lightweight or partial fine-tuning.

We also systematically compared several adaptation strategies for pretrained models, including frozen-feature transfer, low-rank adaptation (LoRA) [41], partial fine-tuning, full fine-tuning, and combinations with knowledge distillation. Partial fine-tuning achieved the best discriminative performance in some experimental settings, whereas LoRA, combined with knowledge distillation, provided a favorable balance among performance, parameter efficiency, and semantic preservation. The interpretability analysis further showed that LoRA with knowledge distillation improved the separability between PE and normal samples while better preserving the semantic structure of the original EchoPrime representation space. By contrast, full fine-tuning enhanced class separation but also induced more pronounced representation drift and weakened the correlation with right-heart-related phenotype readouts derived from the original EchoPrime model. This finding has important methodological implications: in the transfer of medical foundation models, the highest classification performance does not necessarily imply the best clinical interpretability or the most robust semantic transfer. For tasks such as PE-related phenotype recognition, which require an understanding of right heart structure and function, preserving anatomical and physiological semantic information encoded in the pretrained model may help maintain stability across external datasets, low-quality images, and different device environments.

The performance of PE recognition across different ultrasound views also reflects the multimodal manifestation of right heart strain in bedside echocardiography. EchoPE achieved high AUROC values across subxiphoid (SX), parasternal long-axis (PSL), inferior vena cava (IVC), parasternal short-axis (PSS), and apical four-chamber (A4) views, although performance varied across views. The SX and IVC views may provide indirect information related to right heart pressure, volume status, and venous return. The PSL and PSS views facilitate the assessment of right ventricular morphology, septal flattening, and left ventricular compression. The A4 view more directly reflects PE-related findings such as right ventricular enlargement, the right-to-left ventricular size ratio, and McConnell’s sign. The ability of the model to learn complementary information from these different views indicates that PE-related right heart strain does not depend on a single ultrasound sign but rather consists of multiple dynamic structural and hemodynamic manifestations. From a practical emergency-care perspective, this suggests that EchoPE does not require a strictly standardized single view but can instead adapt to the range of bedside views available under different patient conditions.

Interpretability analyses provided further support for the model’s decision-making process. Spatial attribution results showed that, after task-specific adaptation, the model focused more strongly on regions consistent with PE-related right heart strain, including the interventricular septum, the relative morphology of the right and left ventricles, and cardiac chamber boundaries. Temporal occlusion experiments showed that frames in the middle portion of the video had a larger effect on predicted probability, suggesting that the model did not rely solely on static image features but also used relatively stable morphological and motion information across the cardiac cycle. This is particularly relevant for PE-related phenotypes, as abnormal septal motion, impaired right ventricular systolic function, and altered chamber proportions are intrinsically dynamic features.

Another important observation was that, particularly in the pretrained models before task-specific adaptation, some attribution remained distributed over non-cardiac foreground regions, near-field regions, or sector boundaries. This phenomenon may indicate that the pretrained model had already acquired an acquisition-context preference during large-scale pretraining, rather than learning such behavior solely from the HOPE dataset. The fact that the 3D ResNet trained from scratch did not show the same degree of attention to these regions further supports the possibility that such context sensitivity was inherited from the pretrained representation. This issue should be interpreted in the broader context of shortcut learning, in which deep models may exploit predictive but non-causal correlations rather than robust task-relevant features [42]. Our regional occlusion experiments on HOPE did not find direct evidence that the adapted model relied exclusively on such regions for prediction; however, these experiments are insufficient to exclude shortcut learning in the pretrained model. Therefore, before clinical deployment, more rigorous external validation, device-stratified analysis, and additional occlusion experiments are needed to further assess whether the model truly relies on robust anatomical and physiological features.

This study has potential clinical implications. PE is a time-sensitive and potentially life-threatening condition in emergency care. Although CTPA is an important diagnostic test, not all patients can undergo CTPA immediately. As far as we know, the HOPE dataset is the first POCUS PE dataset collected for research purposes, and it has clear value and contributes to the development of echocardiography-based AI models. For patients who are hemodynamically unstable, difficult to transport, or treated in resource-limited environments, bedside ultrasound can rapidly provide information on right heart strain. EchoPE is not intended to replace CTPA or integrated clinical judgment. Rather, it may serve as an assistive tool that provides early risk alerts within the emergency POCUS workflow. For example, when the model identifies PE-related right heart strain phenotypes in bedside videos, it may prompt clinicians to expedite further imaging, intensify monitoring, assess hemodynamic risk, and integrate the findings with clinical presentation, D-dimer levels, myocardial injury biomarkers, and other diagnostic information [7]. In primary-care or resource-limited settings where advanced ultrasound expertise is less available, EchoPE may also help improve the consistency of POCUS interpretation and reduce diagnostic delays caused by variability in operator experience.

### 3.1 Limitations

Several limitations should be acknowledged. First, the labels in HOPE were based on emergency physicians’ interpretation of ultrasound findings, and the main classification task distinguished normal studies from PE-related right heart phenotypes. Although all patients with PE-related right heart phenotypes had CTPA-confirmed PE, the model identifies the PE-related right heart strain phenotype rather than PE itself as defined by CTPA. Some patients with PE may not show obvious right heart strain, whereas some non-PE conditions, such as pulmonary hypertension, chronic lung disease, right ventricular infarction, or abnormal volume loading, may produce similar right heart ultrasound phenotypes. Therefore, the output of EchoPE should be interpreted as a risk signal for PE-related right heart pressure overload, rather than as an independent diagnostic result for PE. Second, this study was primarily based on retrospective real-world POCUS data. Although this design improves the representativeness of the clinical setting, it may still be affected by case selection, scanning indications, operator bias, device differences, and single-center or regional data distributions. Third, although we performed internal testing, view-specific analysis, and interpretability analysis, prospective validation across external institutions, ultrasound devices, operators, and patient populations with different disease spectra is still required to evaluate model generalizability and clinical safety.

In addition, the current model primarily processes a single video as input, whereas real-world emergency POCUS interpretation is often based on multiple views, multiple videos, real-time interactive scanning, and clinical context. Although this study explored cross-view information fusion, simple dual-view fusion or feature hallucination did not yield stable improvements, suggesting that cross-view modeling requires designs that better reflect clinical workflows. Future work may consider patient-level multi-video aggregation, temporal modeling of the scanning process, and multimodal models that incorporate vital signs and laboratory results. Moreover, the current interpretability analysis was mainly based on attention, attribution, and occlusion experiments. These methods provide useful clues regarding model focus, but they cannot fully establish causal mechanisms. Future studies could incorporate more detailed expert annotations and quantitative phenotypic measurements to enable more direct physiological validation of model features.

### 3.2 Conclusion

In conclusion, this study introduced the HOPE dataset and the EchoPE framework, demonstrating the feasibility of adapting a large-scale pretrained cardiac ultrasound foundation model to the emergency recognition of PE-related right heart phenotypes. EchoPE achieved strong classification performance on real-world POCUS videos, and view-specific and interpretability analyses showed that the model can leverage structural and dynamic information related to right heart strain. These findings suggest that task-specific adaptation for acute disease phenotypes, particularly approaches combining partial fine-tuning, parameter-efficient fine-tuning, and knowledge distillation, may represent an important pathway for translating ultrasound foundation models into bedside clinical applications. HOPE and EchoPE provide a generalizable research framework for future development of AI-assisted systems for emergency, low-resource, and operator-heterogeneous environments.

## 4 Acknowledgments

This work was supported by the National Institute of Biomedical Imaging and Bioengineering, National Institutes of Health, under Award Number 5R21EB030677-02. We thank the clinical and technical teams who contributed to data acquisition and curation for the HOPE dataset. We also acknowledge the patients whose data made this research possible.

## 5 Author Contributions

L.Y. (Luming Yang) led the study and was responsible for study design, model development, experimental implementation, data analysis, and manuscript drafting. Y.O. (Yasemin Ozkut) contributed to the early conception of the study and preliminary model development. Z.B. (Zhenyu Bu) conducted the comparative experiments involving foundational echocardiography models and contributed to experimental validation. S.L. (Siqing Li) implemented and evaluated models trained from scratch, contributed to data analysis, and assisted with manuscript writing and figure preparation. C.Y. (Chang Yu) conducted the background literature review, assisted in the experimental process, and verified the foundational datasets. E.S.-L. (Elaine Situ-LaCasse), J.A. (Josie Acuña, M.D.), A.Ya. (Adrienne Yarnish, M.D.), and M.D. (Martin Demant, M.D.) contributed to data collection, curation, and annotation of the dataset. S.R.A. (Srikar R. Adhikari, M.D.) collected the HOPE dataset, provided clinical expertise, secured funding, contributed to study design and interpretation of clinical findings, and supervised the clinical components of the work. A.Y. (Alper Yilmaz, Ph.D.) conceived and supervised the overall project, secured funding, guided the methodological framework and research direction, provided critical feedback on experimental design, and revised and edited the manuscript for intellectual content. All authors reviewed and approved the final manuscript.

## 7 Methods

### 7.1 Study design and ethics

This retrospective study used the HOPE dataset, a de-identified emergency department point-of-care ultrasound dataset developed in collaboration with the University of Arizona. The dataset consisted of archived cardiac POCUS video clips acquired during routine clinical care. All videos were de-identified before transfer to the research team, and no protected health information or directly identifiable patient-level information was available to the investigators.

The study protocol was approved by the University of Arizona Institutional Review Board on May 1, 2023 (IRB Parent Protocol ID: 2003423113). Because the study used retrospective, de-identified data and involved no direct patient contact, the requirement for informed consent was waived by the Institutional Review Board.

### 7.2 Study cohort

The HOPE dataset included adult emergency department patients who underwent bedside cardiac POCUS as part of a routine clinical evaluation. Eligible studies were those containing cardiac or hemodynamic ultrasound videos acquired in the context of suspected pulmonary embolism, PE-related risk, or clinical concern for right-heart strain.

The final analyzable cohort contained 4,069 independent patients, with one video clip per patient. The dataset included 1,934 normal videos and 2,135 videos labeled as PE-related right-heart phenotype. The available ultrasound views included apical four-chamber, parasternal long-axis, parasternal short-axis, inferior vena cava, subxiphoid, and apical four-chamber videos annotated for McConnell’s sign [8] when applicable.

When multiple eligible clips were available for the same patient, only the first eligible clip was retained to minimize data leakage. Because the data were de-identified before transfer, all model development and evaluation were performed at the video level, with each video corresponding to a unique patient.

### 7.3 Ultrasound acquisition

POCUS examinations were performed in the emergency department by clinicians trained in bedside ultrasound, using clinical ultrasound systems available at the participating institution during the study period. The systems included devices from Mindray/Zonare, Philips, GE, and SonoSite. The approximate device distribution followed the institutional archive composition: 45% Mindray/Zonare, 30% Philips, 20% GE, and 5% SonoSite.

The dataset contained views commonly used for the assessment of right-heart strain and hemodynamics: apical four-chamber, parasternal long-axis, parasternal short-axis, inferior vena cava, and subxiphoid views. These views provide complementary information regarding right ventricular size, interventricular septal motion, right ventricular systolic function, ventricular interdependence, and venous congestion.

### 7.4 Reference labels and annotation

All videos were labeled at the clip level. The primary binary label was normal versus PE-related right-heart phenotype. Videos labeled as normal showed no expert-recognized sonographic evidence of PE-related right-heart strain or dysfunction. Videos labeled as PE-related right-heart phenotype demonstrated findings consistent with acute right-heart pressure overload in the setting of confirmed PE, including right ventricular enlargement, an abnormal right ventricular-to-left ventricular size ratio, septal flattening or paradoxical septal motion, impaired right ventricular systolic function, McConnell’s sign [8], or other expert-recognized features of acute right-heart strain.

All PE-positive cases corresponded to patients with PE confirmed by CT pulmonary angiography. Therefore, the PE-related class represented an ultrasound phenotype observed in confirmed PE cases rather than a diagnosis of PE based on ultrasound alone.

Labels were generated by emergency physicians with expertise in POCUS. Each clip was reviewed at the video level and assigned to the corresponding diagnostic category. Cases with diagnostic uncertainty or disagreement were adjudicated through expert consensus. For apical four-chamber videos with sufficient image quality, an additional annotation was performed for McConnell’s sign, defined as regional hypokinesis of the right ventricular free wall with relative preservation of apical motion [8, 43].

No frame-level annotations were used. Clip-level labeling was chosen because bedside echocardiographic interpretation is inherently dynamic, and diagnostically relevant features may appear across multiple frames owing to cardiac motion, respiratory motion, probe motion, and transient changes in image quality.

### 7.5 Video preprocessing

All MP4 videos were decoded into frame sequences and converted to grayscale. If the frame rate exceeded the predefined maximum, frames were uniformly subsampled. To isolate the ultrasound sector, we computed a cross-frame mean-intensity image, thresholded the non-black region, applied morphological opening to remove thin interface artifacts, applied closing to fill small holes, retained the largest connected component, and used its convex hull as the final sector mask. Pixels outside the mask were set to zero. The masked region was cropped to its bounding box, padded to a square canvas, and resized to 256 × 256 pixels by default.

Each video was represented as a tensor:

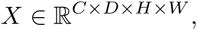

where *C* is the number of channels, *D* is the temporal length, and *H, W* are the spatial dimensions. Pixel values were normalized before model input.

### 7.6 Data splitting and cross-validation

Experiments used patient-level five-fold cross-validation generated from the preprocessed manifest. Only samples with preprocessing status ok were included. Labels were encoded as 0 for normal and 1 for the PE-related phenotype.

Fold generation was stratified by the joint key:

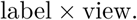

Within each stratum, samples were sorted by identifier, shuffled with split seed 2026, and assigned to five test folds by round-robin allocation. Thus, every patient appeared in exactly one test fold across the full cross-validation.

For each fold, approximately 20% of the cohort was used as the held-out test set. The remaining samples were split into training and validation sets using the same stratification key and a fold-specific seed 2026 + 10000 + *k*, where *k* is the fold index. The target split was approximately 70% training, 10% validation, and 20% testing. Train, validation, and test sets were verified to be mutually exclusive.

### 7.7 Data augmentation

Augmentation was applied only during training. Validation and test videos were evaluated without augmentation.

Intensity augmentation was applied with probability 0.5. When applied, brightness shift, contrast scaling, gamma correction, and Gaussian noise were each independently applied with probability 0.5. Parameters were shared across all frames to preserve temporal consistency. Intensities were clipped to [0, 255].

Spatial affine augmentation was applied with probability 0.8, using rotation within ±15^◦^, scaling from 0.85 to 1.15, and translation up to 15% of the image’s height and width. The same affine grid was applied to all frames.

Temporal cropping was applied with probability 0.3. A contiguous segment of length

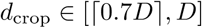

was sampled and interpolated back to a length *D*.

### 7.8 PE classification model

The primary classifier used an EchoPrime video encoder based on MViT-v2-S [44], producing a 512-dimensional video embedding. A binary classification head predicted normal versus PE-related phenotype from this embedding.

We evaluated frozen-encoder training, partial or full fine-tuning, and low-rank adaptation [41]. In LoRA experiments, trainable low-rank adapters were inserted into selected attention projections, primarily QKV and attention-output projections, while pretrained encoder weights remained frozen. Classification heads included multilayer perceptron and residual multilayer perceptron variants. In view-conditioned models, a coarse ultrasound-view representation was concatenated with the video embedding before classification.

### 7.9 Knowledge distillation

Knowledge-distillation [45] experiments used a frozen EchoPrime encoder as the teacher. The student was trained using binary classification loss plus feature-level distillation.

Cosine distillation aligned student and teacher embeddings for the same video:

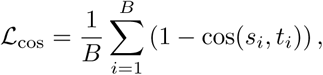

where *s_i_* and *t_i_* are the student and teacher embeddings.

Relational knowledge distillation [46] preserved pairwise batch geometry. Student and teacher pairwise Euclidean distance matrices were normalized by their non-zero mean distances and aligned using Smooth L1 loss [47]:

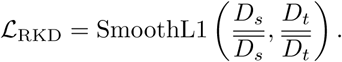

The teacher branch was evaluated without gradient updates. The combined distillation loss was:

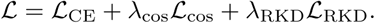

### 7.10 Training and model selection

Each fold was trained independently. Models were trained on the training set, tuned on the validation set, and evaluated once on the held-out test set. Training used class-weighted cross-entropy, weighted random sampling, AdamW optimization [48], linear warm-up, cosine learning-rate decay [49], and mixed precision when CUDA was available. The training seed was set to 2026.

### 7.11 Evaluation Metrics

For each validation and test sample, the model output probability for the PE-related class was saved. Threshold-independent performance was assessed using the area under the receiver operating characteristic curve (AUROC) [50], average precision or area under the precision–recall curve (AUPRC) [51], and the Brier score [52].

Threshold-dependent metrics included accuracy, balanced accuracy, sensitivity, specificity, positive predictive value, negative predictive value, F1 score, and confusion-matrix counts. The primary operating threshold was selected on the validation set by maximizing Youden’s *J* statistic [53]:

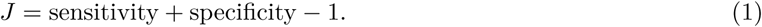

The selected threshold was then fixed and applied to the corresponding test set. In an additional high-sensitivity analysis, we evaluated precision at fixed recall (sensitivity) levels of 0.90 and 0.95, denoted P@R90 and P@R95. For each target recall level, the operating threshold achieving the corresponding recall was selected on the validation set and then fixed before application to the corresponding test set.

For cross-validation reporting, each metric was first computed separately on the held-out test set of each fold. The primary summary across folds was reported as mean ± standard deviation. Pooled out-of-fold predictions were also used to generate aggregate receiver operating characteristic and precision–recall curves.

### 7.12 Statistical Analysis

Uncertainty estimates were computed using non-parametric bootstrap resampling. Unless otherwise specified, 1,000 bootstrap iterations were used to estimate 95% confidence intervals for AUROC, AUPRC, and threshold-dependent metrics. For pooled out-of-fold analyses, resampling was performed at the patient/video level.

When comparing AUROC between models, paired bootstrap testing was used on matched out-of-fold predictions. For threshold-dependent classification metrics, paired bootstrap testing or McNemar’s test [54] was used when appropriate. A two-sided *P <* 0.05 was considered statistically significant. Analyses were performed using Python with PyTorch [55], PyTorch Lightning, scikit-learn [56], NumPy, pandas, and SciPy. Exact package versions are reported in the codebase provided in section 9.

### 7.13 View-Classification Experiments

A separate set of experiments evaluated ultrasound view classification. These experiments used the ConvNeXt-base view classifier [57] and EchoPrime parameters as initialization if specified as pretrained. Only the first preprocessed frame from each video was used for view recognition.

The evaluated strategies included the direct use of the pretrained classifier, linear probing with a newly initialized classification head, partial fine-tuning of the final ConvNeXt stage and head, full fine-tuning, and training from random initialization. Data-scaling experiments used training fractions of 0.15, 0.30, 0.50, 0.75, and 1.00 across three random seeds.

View-classification performance was summarized using accuracy, balanced accuracy, macro F1 score, weighted F1 score, per-class recall, mean one-versus-rest accuracy, and row-normalized confusion matrices.

### 7.14 Interpretability Analysis

Interpretability analyses were performed on selected checkpoints from the PE classification experiments. For each selected model, sample-level predictions, 512-dimensional embeddings, attention-derived maps, gradient-based attribution maps, and temporal occlusion curves were saved.

Spatial attribution was computed using gradient-times-input maps and MViT attention aggregation. Attention maps were generated by aggregating class-token-to-patch attention from the latter half of the MViT blocks and upsampling the resulting patch-level map to the input video resolution. Temporal gradient sensitivity was computed by summing positive attribution values over the spatial dimensions of each frame.

Temporal occlusion was used as the primary temporal evidence analysis. Contiguous windows of frames were set to zero, and the change in PE probability or PE logit relative to the original prediction was recorded. Larger decreases after occlusion were interpreted as greater model reliance on the occluded temporal segment.

### 7.15 Representation Analysis

Model representations were analyzed using the 512-dimensional video embeddings. Dimensionality-reduction analyses included principal component analysis (PCA) [58], UMAP [59], and t-SNE [60]. Linear probes were trained to assess the extent to which PE labels and ultrasound view labels were encoded in the learned representations. Label and view separation were quantified using silhouette scores.

Embedding drift was measured by computing the cosine similarity between embeddings from fine-tuned models and embeddings from the frozen EchoPrime encoder for the same videos. Semantic preservation was additionally evaluated by passing frozen and fine-tuned embeddings through EchoPrime phenotype or report probes when available and comparing the resulting phenotype scores using Spearman correlation.

### 7.16 ROI and Perturbation Analysis

For selected cases, weakly supervised acquisition-context masks were generated from the preprocessed videos. Four automatic ROI categories were used: foreground ultrasound sector, background, sector border, and probe near-field. The foreground sector was estimated from mean image intensity across channels and time. The sector border was defined using dilation and erosion of the foreground mask. The probe near-field was defined as the superior portion of the foreground sector.

Optional anatomical ROI annotations were supported using polygon masks for the right ventricle, left ventricle, interventricular septum, D-sign region, and the right ventricle–left ventricle boundary.

For each attribution map and ROI, attribution mass, ROI area fraction, ROI enrichment, and overlap with the top 10% highest-attribution pixels were computed. To evaluate regional sensitivity, each ROI was spatially occluded across all frames using zero filling in normalized video space. The change in PE probability and PE logit relative to the unoccluded prediction was measured and summarized by model, view, true label, attribution type, and ROI category.

### 7.17 Data Protection and Compliance

All videos were de-identified before transfer to the research team. The research team did not receive patient names, medical record numbers, dates of birth, accession numbers, full examination dates, or other direct identifiers. De-identified videos were stored in secure institutional cloud storage managed by the participating institution. Data sharing between the clinical site and the research team occurred through institutionally approved secure platforms. All analyses were conducted only on de-identified video clips and aggregate cohort-level data.

## 8 Data Availability

The HOPE dataset is subject to institutional and ethical regulations. De-identified data will be made publicly available upon article acceptance through an appropriate public repository, in accordance with institutional review board (IRB) policies and applicable data-sharing agreements. Details and access instructions will be provided at the time of publication.

## 9 Code Availability

The appendix (supplementary material) and all code used in this study, including data preprocessing pipelines, model training scripts, analysis workflows, and experimental configurations, will be made available upon article acceptance.

## Footnotes

1 The appendix (supplementary material) and code will be released upon acceptance.

